# Implementation of an ERAS program in patients undergoing thoracic surgery at a third-level university hospital. An ambispective cohort study

**DOI:** 10.1101/2020.10.07.20197962

**Authors:** Soledad Bellas-Cotán, Rubén Casans-Francés, Cristina Ibáñez, Ignacio Muguruza, Luis E. Muñoz-Alameda

## Abstract

**Objetive:** *T*o analyze the effects of the implementation of an ERAS program in patients undergoing pulmonary resection in a tertiary university hospital on the rates of complications and readmission and the length of stay.

**Methods:** ambispective cohort study, with a prospective arm of patients undergoing thoracic surgery within an ERAS program versus a retrospective arm of patients before the implementation of the protocol. We recluited 50 patients per arm. The primary outcome was the number of patients with 30-day surgical complications. Secondary outcome included ERAS adherence, no-surgical complications, mortality, readmission, reintervention rates, pain and hospital lenght of stay. We performed a multivariate logistic analysis to study the association of coutcomes with ERAS adherence.

**Results:** We found no difference between the two groups in surgical complications [Standard 18 (36%) vs 12 (24%], p =0.19]. ERAS group was significantly lower only in its readmission rate [Standard 15 (30%) vs 6 (12%], p =0.03]. In multivariate analyses, ERAS adherence was the only factor associated with a reduction in surgical complications [OR (95%CI) = 0.02 (0.00, 0.59), p = 0.03] and length of stay [HR (95%CI) = 18.5 (4.39, 78.4), p < 0.001].

**Conclusions:** ERAS program was able to decrease the readmission rate at our centre significantly. The adherence to the ERAS protocol influenced the reduction of surgical complications and length of stay.

## INTRODUCTION

Lung cancer is the leading cause of cancer death worldwide, representing 20,55% and 14% of cancer deaths in Spain^1^ and the United States^2^, respectively. Currently, pulmonary resection is the treatment of choice for lung cancer^3^. However, this surgery is associated with significant complications in almost 50% of the cases, possibly delaying patient recovery and consequently increasing hospitalisation costs^4^.

Professor Henrik Kehlet described ERAS programs at the end of the last century^5^. His ideas were that the application of specific measures based on scientific evidence during the perioperative period of the patient could decrease the stress produced by surgical aggression^6^. Thus, in recent years, ERAS programs have proven effective in reducing surgical complications, length of stay and hospital costs^7–9^.

Over the last years, specific ERAS surgical approaches have been described for thoracic surgery^10–12^. Nevertheless, there is still a lack of evidence to support ERAS programs for pulmonary resection surgery, particularly in terms of clinical results combined with minimally invasive procedures.

Our study aims to analyze the effects of the implementation of an ERAS program in patients undergoing pulmonary resection in a tertiary university hospital on the rates of complications and readmission and the length of stay.

## METHODS

### Study design and participants

This study analyzes the implementation of an ERAS program in a thoracic service of a third level hospital *(Hospital Fundación Jiménez Díaz, Madrid, Spain)*. To this end, we designed an ambispective cohort study, with a prospective arm of patients undergoing thoracic surgery within an ERAS program versus a retrospective arm of patients before the implementation of the protocol. Our centre’s ethics committee approved our study before the start of patient recruitment, January 2018 Ref: EO071-18_FJD and has been registered in the database of clinical studies ClinicalTrials.gov Identifier: NCT04579601. This study followed the Strengthening the Reporting of Observational Studies in Epidemiology (<u>STROBE</u>) reporting guideline for cohort studies^13^.

After informed consent, we included patients consecutively since the implementation, except those who refused the inclusion in the study or were under 18 years old. We also asked for informed consent from the patients who were part of the retrospective cohort. For the calculation of the required sample size, we assumed that the ERAS program would result in a 25% reduction in the absolute risk of suffering a surgical complication. Since the surgical complication rate for our patients in 2016 was 40%, a type-I error of 5% and a power of 80% would require 47 patients per arm.

### Procedures

We recruited 50 patients throughout 2018 and 2019 and compared them with data from the last 50 patients in 2016, the year in which we knew the surgical complication rate. We followed up each patient for 30 days after surgery through hospital and primary care medical records. Demographic and comorbidity data were collected from all patients, from which we calculated Charlson’s comorbidity index^14^ for all patients.

We designed our centre’s ERAS program through different measures during the preoperative, intraoperative and postoperative period. During the preoperative period, the patients and their families received comprehensive multidisciplinary information about the protocol, as well as their daily goals and expected discharge date. Also, a team specialized in therapy against lung diseases taught patients pulmonary expansion exercises to be carried out until surgery was performed. Smoking cessation and nutritional screening of the patient were also part of this stage.

The patients underwent video-assisted thoracoscopic surgery (VATS), whenever possible, leaving a chest tube at the end of the surgery. All subjects received antibiotic and antithrombotic prophylaxis. Intraoperative management of patients was performed under general anaesthesia combined with regional techniques for pain control, avoiding the use of benzodiazepines and opioids. Those patients in whom thoracic epidural catheter was implemented during surgery continued its use through a patient-controlled analgesia system. Besides, a hot air system warmed the patients during surgery to maintain normothermia. Extubation was performed as soon as possible after the end of the surgery, and we encouraged early removal of the urinary catheter.

From the time of extubation, the patients began oral tolerance and respiratory physiotherapy exercises. Also, the patients were allowed to walk around early. The patients were discharged when they were free of complications, without severe pain, urinary catheter or chest tube.

### Outcomes

The primary outcome was the number of patients with 30-day surgical complications. We defined air leakage, bleeding, infection, and reintervention as surgical complications. Secondary outcome included ERAS adherence, no-surgical complications, mortality, readmission, reintervention rates, pain (defined as any level of pain that prevents early ambulation) and hospital lenght of stay. To evaluate ERAS adherence, we defined seven items: VATS approach, regional analgesia, oral tolerance within 6 hours, urinary catheter removal within 24 hours, ambulation within 24 hours, respiratory physiotherapy within 24 hours and chest tube removal within 48 hours.

### Statistical analysis

We analyzed outcomes depending on whether the patient belonged to the ERAS program or the retrospective standard cohort. The discrete and continuous variables were described as number and percentage and median (interquartile range [IQR]) and their differences analyzed using the Pearson test or the Wilcoxon rank-sum tests. Subsequently, according to the adherence rate to ERAS items (regardless of whether the patients belonged to the ERAS or the retrospective standard cohort), We performed a multivariate logistic analysis to study the association of complication rates, readmission or pain with ERAS adherence, clinical and demographic data, presenting the results in forest plots as odds ratio with 95% confidence interval. Similarly, we used Cox regression for multivariate analysis of length of stay, presenting the results in forest plot as hazard ratio with 95% confidence interval. To avoid errors by multiple comparisons, we calculated the respective q-value for each p-value to maintain a false discovery rate below 5%^15^. We considered comparisons in which p-value and q-value were below .05 as being statistically significant.

## RESULTS

No patient declined inclusion in the study. The demographic characteristics and comorbidities of the patients are shown in Table 1. The two cohorts were not totally homogeneous, with a higher number of patients with hypertension in the standard cohort [26 (52%) vs ERAS 15 (30%), p = 0.03] and chronic obstructive pulmonary disease [12 (24%) vs ERAS 4 (8%), p = 0.02]. Although the number of patients with ASA class > 2 was higher in the standard group [26 (52%) vs ERAS 15 (30%)], we found no difference between the cohorts in Charlson’s comorbidity index. We included these three items, along with age and sex in the subsequent multivariate analyses.

**TABLE 1.**
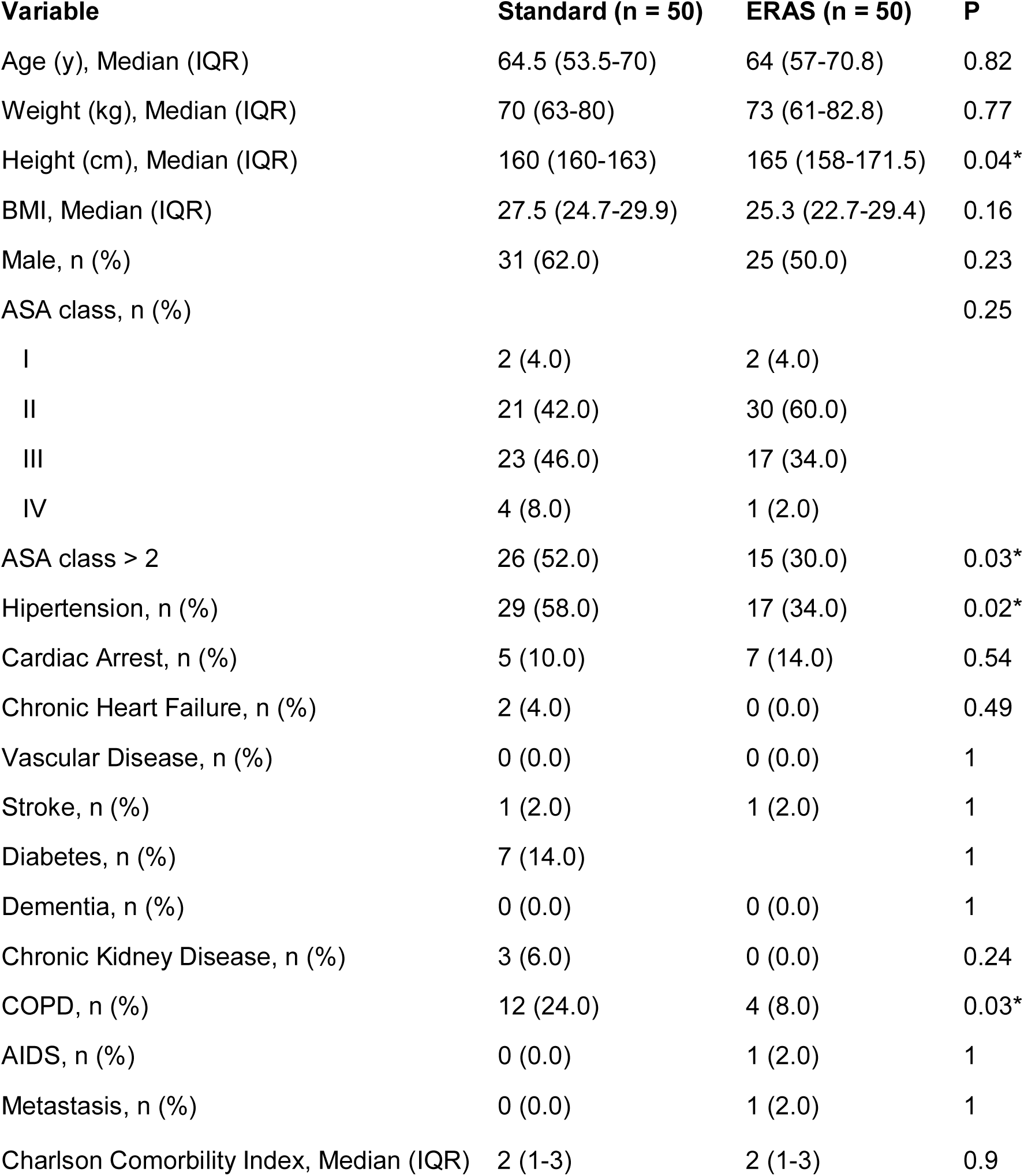
Patient demographics and comorbidity data. Pearson or Wilcoxon tests were applied depending on whether the variable was discrete or continuous. It is considered significant p < 0.05.

| **Variable** | **Standard (n = 50)** | **ERAS (n = 50)** | **P** |
| --- | --- | --- | --- |
| Age (y), Median (IQR) | 64.5 (53.5-70) | 64 (57-70.8) | 0.82 |
| Weight (kg), Median (IQR) | 70 (63-80) | 73 (61-82.8) | 0.77 |
| Height (cm), Median (IQR) | 160 (160-163) | 165 (158-171.5) | 0.04* |
| BMI, Median (IQR) | 27.5 (24.7-29.9) | 25.3 (22.7-29.4) | 0.16 |
| Male, n (%) | 31 (62.0) | 25 (50.0) | 0.23 |
| ASA class, n (%) |  |  | 0.25 |
| I | 2 (4.0) | 2 (4.0) |  |
| II | 21 (42.0) | 30 (60.0) |  |
| III | 23 (46.0) | 17 (34.0) |  |
| IV | 4 (8.0) | 1 (2.0) |  |
| ASA class > 2 | 26 (52.0) | 15 (30.0) | 0.03* |
| Hipertension, n (%) | 29 (58.0) | 17 (34.0) | 0.02* |
| Cardiac Arrest, n (%) | 5 (10.0) | 7 (14.0) | 0.54 |
| Chronic Heart Failure, n (%) | 2 (4.0) | 0 (0.0) | 0.49 |
| Vascular Disease, n (%) | 0 (0.0) | 0 (0.0) | 1 |
| Stroke, n (%) | 1 (2.0) | 1 (2.0) | 1 |
| Diabetes, n (%) | 7 (14.0) |  | 1 |
| Dementia, n (%) | 0 (0.0) | 0 (0.0) | 1 |
| Chronic Kidney Disease, n (%) | 3 (6.0) | 0 (0.0) | 0.24 |
| COPD, n (%) | 12 (24.0) | 4 (8.0) | 0.03* |
| AIDS, n (%) | 0 (0.0) | 1 (2.0) | 1 |
| Metastasis, n (%) | 0 (0.0) | 1 (2.0) | 1 |
| Charlson Comorbidity Index, Median (IQR) | 2 (1-3) | 2 (1-3) | 0.9 |

Data on ERAS adherence and compliance for each of the protocol items are shown in Table 2. Adherence to the ERAS protocol was significantly higher in the prospective than in the retrospective cohort [Median: Standard 0.29 (0.14-0.43) vs ERAS 0.71 (0.57-0.82), p < 0.001]. The VATS approach was greater in the ERAS group [29 (58%) vs Standard 11 (22%), p < 0.001], and the number of patients who ambulated on the first postoperative day [40 (80%) vs Standard 0 (0%), p < 0.001], but no difference was found in the use of regional analgesia. Also, the times to oral intake and removal of the urethral catheter were lower in the ERAS group [Median (h): Standard 24 (24-24) vs ERAS (6-7.5), and Standard 48 (24-48) vs ERAS 19 (6-24), respectively).

**TABLE 2.**
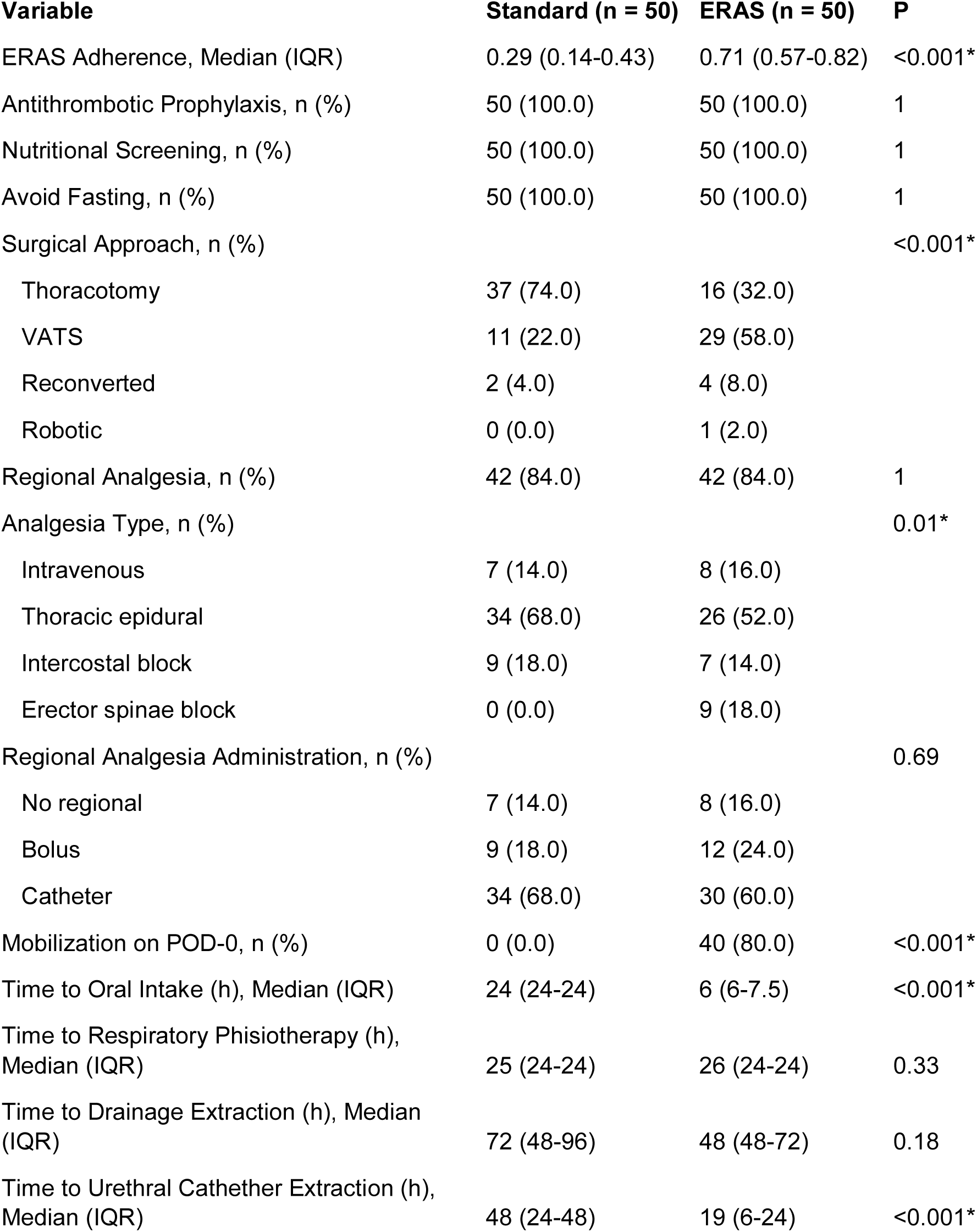
ERAS adherence data. Pearson or Wilcoxon tests were applied depending on whether the variable was discrete or continuous. It is considered significant p < 0.05.

| **Variable** | **Standard (n = 50)** | **ERAS (n = 50)** | **P** |
| --- | --- | --- | --- |
| ERAS Adherence, Median (IQR) | 0.29 (0.14-0.43) | 0.71 (0.57-0.82) | <0.001* |
| Antithrombotic Prophylaxis, n (%) | 50 (100.0) | 50 (100.0) | 1 |
| Nutritional Screening, n (%) | 50 (100.0) | 50 (100.0) | 1 |
| Avoid Fasting, n (%) | 50 (100.0) | 50 (100.0) | 1 |
| Surgical Approach, n (%) |  |  | <0.001* |
| Thoracotomy | 37 (74.0) | 16 (32.0) |  |
| VATS | 11 (22.0) | 29 (58.0) |  |
| Reconverted | 2 (4.0) | 4 (8.0) |  |
| Robotic | 0 (0.0) | 1 (2.0) |  |
| Regional Analgesia, n (%) | 42 (84.0) | 42 (84.0) | 1 |
| Analgesia Type, n (%) |  |  | 0.01* |
| Intravenous | 7 (14.0) | 8 (16.0) |  |
| Thoracic epidural | 34 (68.0) | 26 (52.0) |  |
| Intercostal block | 9 (18.0) | 7 (14.0) |  |
| Erector spinae block | 0 (0.0) | 9 (18.0) |  |
| Regional Analgesia Administration, n (%) |  |  | 0.69 |
| No regional | 7 (14.0) | 8 (16.0) |  |
| Bolus | 9 (18.0) | 12 (24.0) |  |
| Catheter | 34 (68.0) | 30 (60.0) |  |
| Mobilization on POD-0, n (%) | 0 (0.0) | 40 (80.0) | <0.001* |
| Time to Oral Intake (h), Median (IQR) | 24 (24-24) | 6 (6-7.5) | <0.001* |
| Time to Respiratory Physiotherapy (h), Median (IQR) | 25 (24-24) | 26 (24-24) | 0.33 |
| Time to Drainage Extraction (h), Median (IQR) | 72 (48-96) | 48 (48-72) | 0.18 |
| Time to Urethral Catheter Extraction (h), Median (IQR) | 48 (24-48) | 19 (6-24) | <0.001* |

The primary and secondary results are shown in Table 3. We found no difference between the two groups in either surgical complications [Standard 18 (36%) vs 12 (24%], p =0.19], non-surgical complications [Standard 21 (42%) vs 12 (24%], p =0.06] or length of stay [Median (days): Standard 4 (3-6) vs 4 (3-5], p =0.19], and the ERAS group was significantly lower only in its readmission rate [Standard 15 (30%) vs 6 (12%], p =0.03]. No deaths were recorded in the ERAS group, compared to two deaths in the retrospective cohort.

**TABLE 3.** Results of main and secondary outcomes. Pearson or Wilcoxon tests were applied depending on whether the variable was discrete or continuous. It is considered significant p < 0.05.

| **Variable** | **Standard (n = 50)** | **ERAS (n = 50)** | **P** |
| --- | --- | --- | --- |
| Surgical Complications, n (%) | 18 (36.0) | 12 (24.0) | 0.19 |
| Other Complications, n (%) | 21 (42.0) | 12 (24.0) | 0.06 |
| Mortality, n (%) | 2 (4.0) | 0 (0.0) | 0.49 |
| Reintervention, n (%) | 2 (4.0) | 0 (0.0) | 0.49 |
| Readmission, n (%) | 15 (30.0) | 6 (12.0) | 0.03* |
| ICU Readmission, n (%) | 0 (0.0) | 0 (0.0) | 1 |
| Pain, n (%) | 16 (32.0) | 10 (20.0) | 0.17 |
| Death, n (%) | 2 (4.0) | 0 (0.0) | 0.49 |
| Length of stay (d), Median (IQR) | 4 (3-6) | 4 (3-5) | 0.39 |

Multivariate analyses are shown in Figures 1, 2 and 3. ERAS adherence was the only factor associated with a reduction in surgical complications [OR (95%CI) = 0.02 (0.00, 0.59), p = 0.03, Figure 1A], and postoperative pain [OR (95%CI) = 0.01 (0.00, 0.28), p = 0.01, Figure 2B]. It was also associated with a lower readmission rate [OR (95%CI) = 0.01 (0.00, 0.24), p = 0.007, Figure 2A] and an increased likelihood of discharge from the hospital [HR (95%CI) = 18.5 (4.39, 78.4), p < 0.001, Figure 3]. The thoracic epidural analgesia was the only factor that showed an association with lower rates of non-surgical complications [OR (95%CI) = 0.09 (0.01, 0.49), p = 0.008, Figure 1B]. It was also associated with lower rates of postoperative pain [OR (95%CI) = 0.16 (0.03, 0.86), p = 0.03, Figure 2A] and increased probability of discharge from the hospital [HR (95%CI) = 3.14 (1.39, 7.07), p = 0.006, Figure 3]. The intercostal blockade also increased the latter likelihood [HR (95%CI) = 7.55 (2.94, 19.3), p < 0.001, Figure 3].

**Figure 1.**
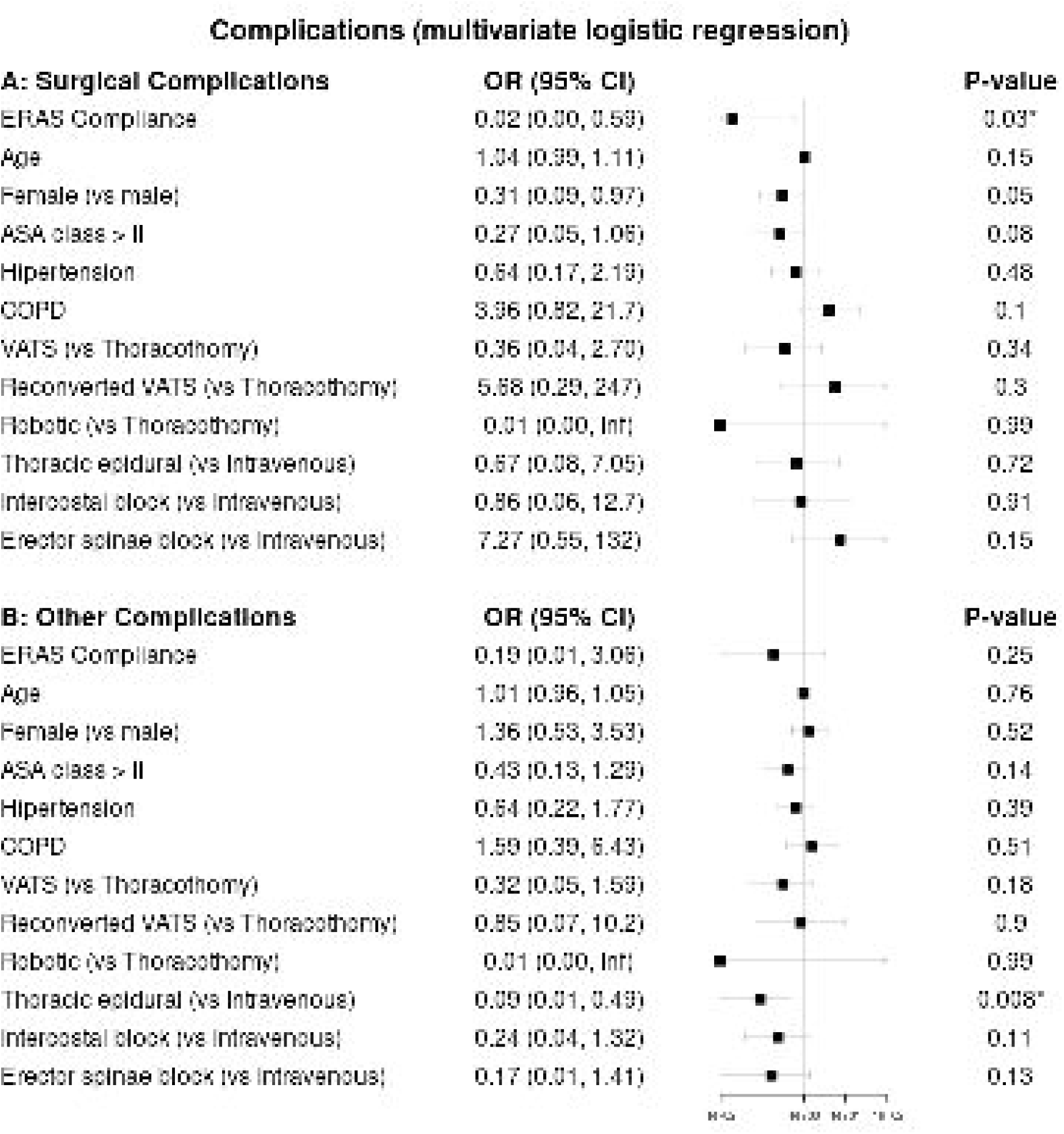
Forest plot of multivariate logistic analysis of the influence of patient comorbidity and ERAS on complications. A: Surgical complications. B: Non-surgical complications. We present the results as an odds ratio with a 95% confidence interval. Results less than 1, left of the y-axis, imply risk reduction. We accept p < 0.05 as significant.

**Figure 2.**
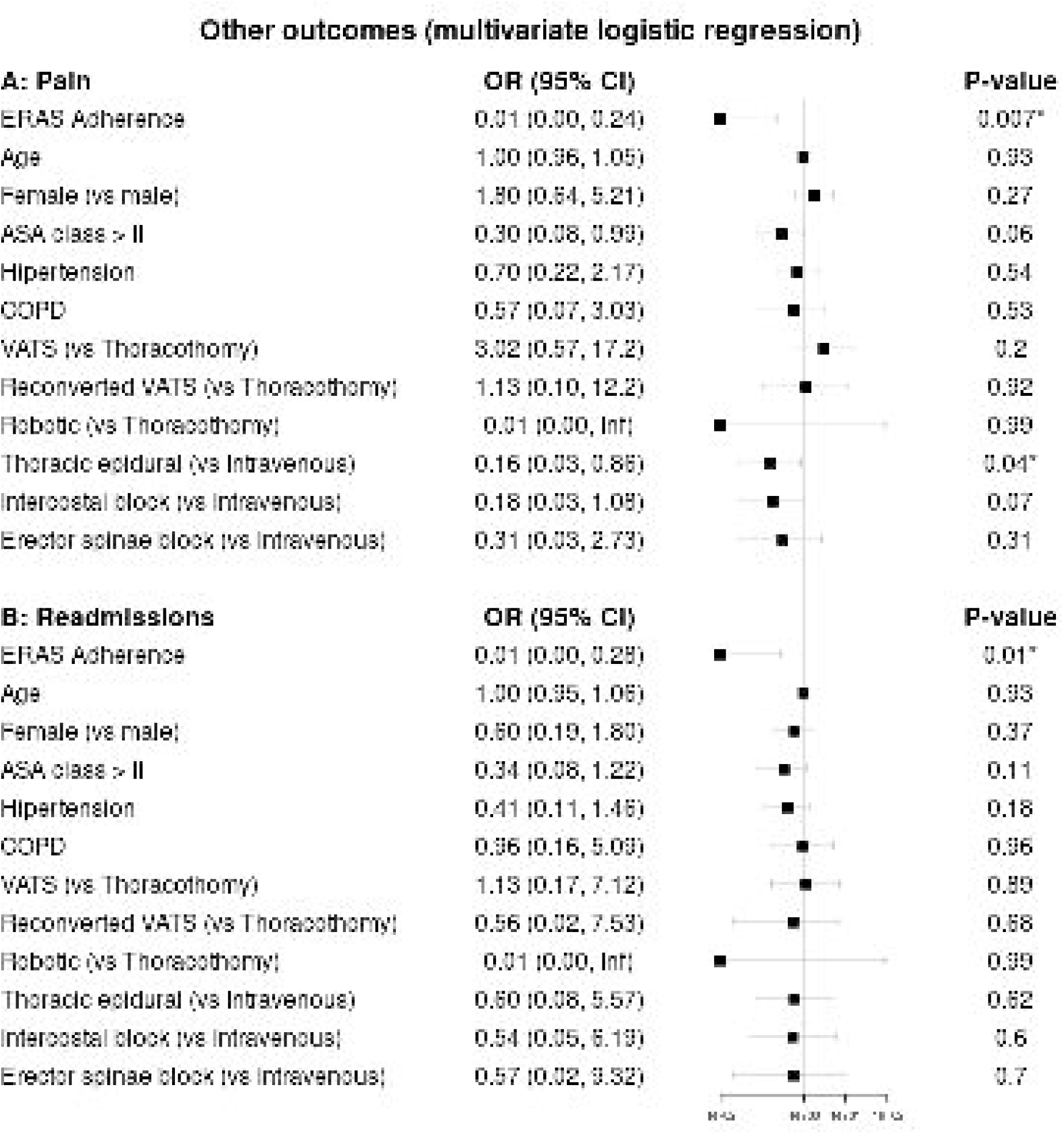
Forest plot of multivariate logistic analysis of the influence of patient comorbidity and ERAS on other outcomes. A: Pain. B: Readmission. We present the results as an odds ratio with a 95% confidence interval. Results less than 1, left of the y-axis, imply risk reduction. We accept p < 0.05 as significant.

**Figure 3.**
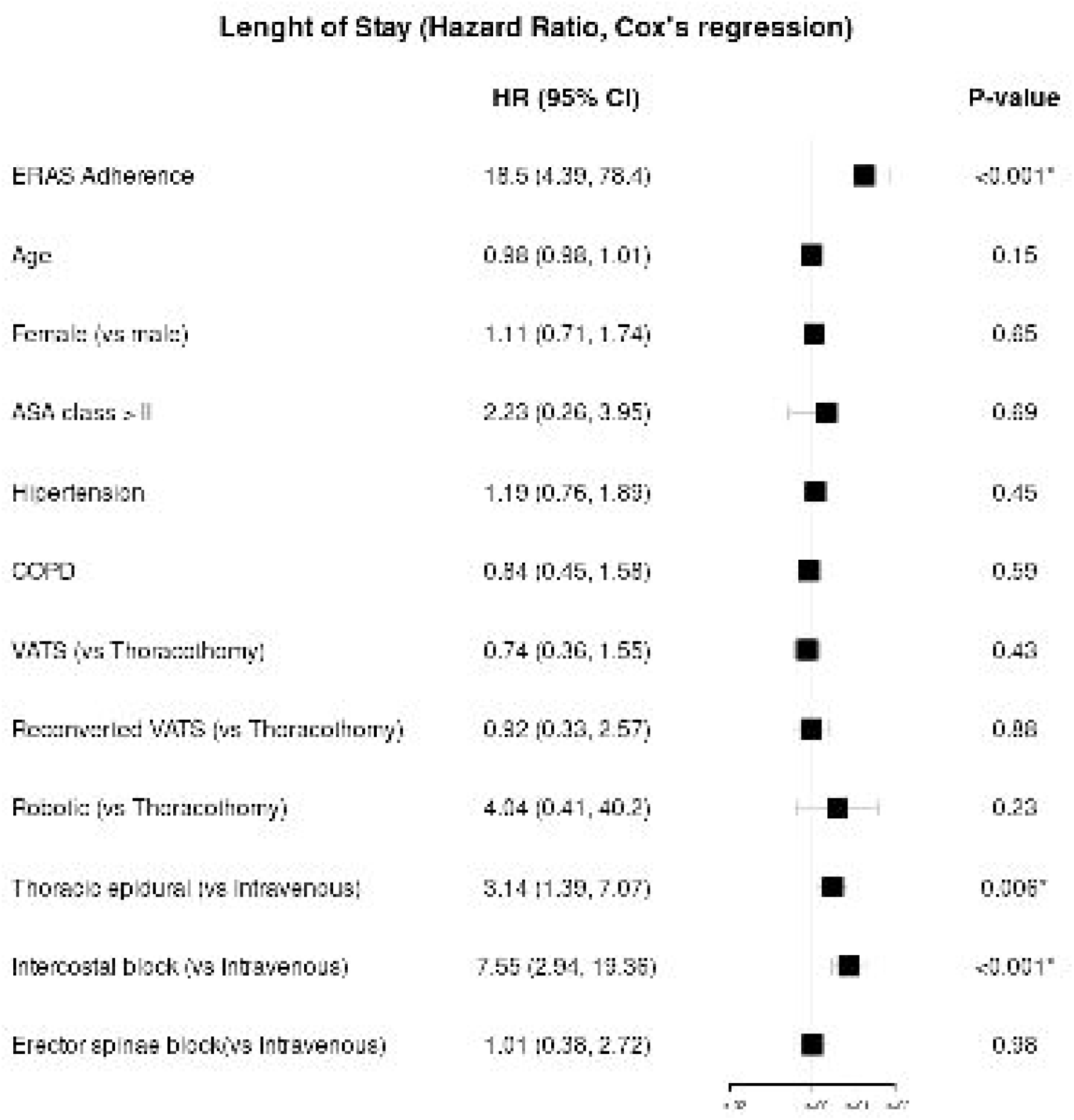
Forest plot of Cox regression of the influence of patient comorbidity and ERAS on length of stay. We present the results as an hazard ratio with a 95% confidence interval. Results more than 1, right of the y-axis, imply increased probability of hospital discharge. We accept p < 0.05 as significant.

No significant p-value was rejected after the calculation of q-value within the multiple comparability study.

## DISCUSSION

Our study has shown how the degree of adherence with the ERAS protocol is one of the essential points towards the reduction of complications in patients undergoing pulmonary resection. Thus, the increase in the degree of adherence was associated with a decrease in complications and length of stay. In other words, while small marginal gains caused by each of the items in isolation show little effect on the results, it is the set of all the measures applied that are responsible for the improvement. These findings are consistent with those of Madani et al.^10^, who observed how compliance with the full ERAS program is probably the most critical factor, more than individually applied elements.

However, it is also important to note that it is one thing to include a patient in an ERAS program and another to have the patient comply with all the items in that program. Resistance to change, especially in the initial phases of the program, can make it challenging to implement the program^16^. If we add to this the fact that there is often a lack of multidisciplinarity on the part of the professionals and that it is not easy to get the patient out of his passive role, we obtain apparent ineffectiveness of the ERAS program. A clear example in our country was the POWER study^17^, a prospective multi-centre study of 80 centres and more than 2000 patients in which the centre declared whether or not it performed ERAS programs on its patients and then independently collected the adherence to such programs. The results of the study showed that although there were no differences in moderate or severe complications between the two groups, there were differences if the patients were divided into adherence quartiles.

One of the critical points of the ERAS programs is that it facilitates standardisation in perioperative patient care. This standardisation tends to translate into improved outcomes, and several studies show it. Cerfolio et al.^18^ implemented an ERAS program for pulmonary resections with an approach focused on the education of the patient, the use of epidural analgesia, the standardised and early withdrawal of urethral catheters and chest drains and early ambulatory. These interventions enabled early discharges without negatively affecting morbidity or mortality. Another example is the randomised controlled clinical trial conducted by Muehling et al.^19^, who observed how a protocol based on avoidance of extended fasting, regional analgesia, early oral feeding tolerance, and early ambulation resulted in a significant reduction in pulmonary complications.

On the other hand, we must also be clear that the fact that it is the sum of the marginal effects of each factor that is responsible for the benefit of the ERAS programs does not mean that the influence of each factor is the same, but that there are more influential factors than others. At this time, the literature does not establish whether it is the adherence to the program or the weight of specific points individually that is responsible for the improvement in thoracic surgery. However, experience in colorectal surgery indicates that both approaches may be correct^20^.

We only found the influence of regional analgesia in the improvement of results, and exclusively in multivariate analyses. This factor has a vital influence on other areas of the protocol since pain is responsible, like immobility. Immobility after thoracic surgery is not uncommon due to factors such as pain, nausea or keeping the chest tube in place^21^. The early mobilisation is a critical factor in the reduction of complications in ERAS programs^22^, but we were unable to find this influence in our study. Another influential factor in early mobilisation is the early removal of chest tubes. Although our study does not show any influence either, the literature shows that the more aggressive the early removal, the better the results^23,24^.

Our study also found no effect of VATS on the results. Minimally invasive surgery seems to be an independent predictor of favourable outcomes after colorectal cancer surgery in ERAS programs^25^. Along these lines, pulmonary resection surgery through VATS has gradually increased its popularity in the light of the new data regarding its effectiveness while results are potentially improved^26,27^.

The feared consequence of implementing an ERAS program for pulmonary resection surgery is the increase in readmissions; this is associated to a reduction of survival in both the short and long term^28^. Throughout the implementation of the ERAS program for pulmonary resection an increase in readmissions has not been found^10^. In our study, the readmission rate was 12%, and it was the only outcome where mere inclusion in the ERAS program was favourable, consistent with other published studies^28^.

Our study found no relationship with performance improvements in certain items of the ERAS protocol. We believe that the main limitation of our study causes this lack of significant findings, that we were too optimistic about the theoretical effect of the ERAS protocol on the complication rate. The estimated value of a 25% absolute risk reduction resulted in a reduction in the theoretical sample size and a decrease in power. A more conservative value would have increased the power of the study.

In conclusion, the ERAS program was able to decrease the readmission rate at our centre significantly. Likewise, the adherence to the ERAS protocol influenced the reduction of surgical complications and length of stay.

## Data Availability

the Author authorized to include all the content of the work, all data and note links.

